# After Death Experiences in Patients with Parkinson’s Disease

**DOI:** 10.1101/2025.08.21.25333962

**Authors:** Courtney Applewhite, Jevita Potheegadoo, Fosco Bernasconi, Olaf Blanke

## Abstract

Hallucinations in Parkinson’s disease (PD) are one of the most common non-motor symptoms, affecting about half of all patients, and the most common form are presence hallucinations (PH). Some individuals with PD report sensing the presence of deceased loved ones, referred to as grief or bereavement hallucinations. In these experiences, people report feeling the presence, hearing, seeing, or otherwise perceiving a deceased person. We refer to them as “after death experiences” (ADEs). We analyzed the phenomenology of ADEs in six patients with PD. Relying on in-depth interviews, we describe the appearance and evolution of ADEs, their emotional aspects, how ADEs manifest, and how patients accounted for the experience. We report several similarities between ADEs in patients with PD and those in bereaved individuals (not related to PD), but also important differences. Patients with PD experience repeated ADEs. This reoccurrence allowed us to examine the way in which people make sense of these experiences by using both cognitive and reflective language. Our findings are not only of relevance for ADEs in PD, but also for a better understanding of ADEs in other contexts, allowing to form new hypotheses about ADEs and their social-cognitive mechanisms to inspire future behavioral and neurocognitive research.

## Introduction

Monique, once a psychiatrist, was interested in and no stranger to perceiving unusual happenings in her work and, lately, in her head. She is in her 70s and has had Parkinson’s disease (PD) for nine years. As her disease has progressed, Monique has experienced many types of hallucinations, in which she has sensed the presence of her close friend or mother when they were not there (both alive), but also of her long-deceased husband. For the latter, she explains that her husband is not “really” with her, but the impression that he is there gives her comfort, nonetheless. We asked her if she was seeing her husband, or his ghost or spirit, and she replied, “I think [the idea] is really pleasant…[but] he is dead.” These kinds of hallucinatory experiences in which the person has a vivid sensation that someone is nearby when no one is actually there are generally called presence hallucinations (or PH), and they are a common symptom of PD (Bernasconi et al., 2021; Fénelon et al., 2011; Ffytche et al., 2017a; Potheegadoo et al., 2021). Outside of this clinical context, when these hallucinations are identified as deceased people, they have often been called grief or bereavement hallucinations (Castelnovo et al., 2015; Grimby, 1993; Kamp et al., 2020). To bridge the terminology between the clinical, often pathologized, term “hallucination” and the more colloquial descriptions of sense of presence, we will call these experiences “after death experiences” (ADEs). People with PH can have PH that may or may not be ADEs.

### ADEs

Between 30-60% of the adult population worldwide has experienced an ADE, ranging from experiences of the sense of presence, as described above, to visual, tactile, auditory, or even olfactory or gustatory experiences of the deceased (Castelnovo et al., 2015). Despite this wide variety of presentation and relatively high prevalence, the research into ADEs is limited. As psychological interest in grief increased throughout the twentieth century, Sigmund Freud and psychoanalysis argued that these experiences reflect the difficulty in severing attachment to the deceased (Freud, 1917). Freud described these experiences as pathological, and something to be overcome through “grief work.” However, by the mid-twentieth century, psychology began to recognize ADEs as temporary and natural parts of the grieving experience rather than necessarily pathological, and were challenging assumptions that these experiences were abnormal (Lindemann, 1944; Miles & Demi, 1984).

Several psychological theories have been proposed to account for these kinds of bereavement experiences and many relate to the social disruptions that occur when someone close to us dies. In attachment theory, grief is a natural response to separation and ADEs may be a result of increased sense of seeking that arises when a loss occurs (Bowlby, 1969; O’Mahony et al., 1984; Parkes, 1970). This underlying attachment theory led to the development of a theory of continuing bonds in which maintaining a relationship with the deceased can be beneficial (Chan et al., 2005; Costello & Kendrick, 2000; Klass et al., 1996). Researchers who argue for a theory of continuing bonds assert that ADEs are a natural consequence of an ongoing relationship with the deceased that physical death does not automatically end.

More recently, perhaps as an extension of these early attachment theories, evolutionary psychological approaches have posited that forms of ADEs may be due to errors in perception when seeking the deceased person in the environment, driven by a survival instinct to keep the group together (White & Fessler, 2018). From an evolutionary perspective, it was adaptive for our ancestors to seek out members of their immediate group or signs of their return after a perilous endeavor like a hunting trip. Such seeking behavior has been argued to have adaptive value in the context of prolonged absences of close kin, but maladaptive in the context of bereavement (Archer, 2001). This and other evolutionary psychological approaches draw our attention to the way in which our behavior and responses to the environment are informed from a group-level perspective across time and cultures as opposed to centering on individual mechanisms.

Alongside these relationship-based explanations, ADEs have also been explained in terms of meaning creation and sense-making around death. The bereaved need to “make sense” of a loss by explaining why it happened in terms of consistent or changing worldviews (Janoff-Bulman, 1992; Park, 2010; Park & Folkman, 1997; Stroebe & Schut, 2010). ADEs occur and the recognition of the experience and the way in which it is understood by the bereaved are part of the way in which the bereaved makes meaning. There have also been shifts from meaning-making based on internal, psychological compulsion to meaning as socially and culturally shaped (Valentine, 2019).

The meaning-making approach and social context help us better understand ADEs when they do not occur just after a death. ADEs are most common in the days and weeks after the death of a close loved one (Grimby, 1993), but they may also occur months or even years later (Rees, 1971). After the acute period of grief, it is less likely that mechanisms related to seeking behavior or attachment are going to be as relevant. Later appearances of ADEs are more difficult to explain, as the theories related to attachment theory, continuing bonds, and seeking behaviors rely on incorrect perceptual mechanisms indicating the deceased person is still alive. However, under the assumption that grief is not a linear process, it is possible that these altered mechanisms and continued ‘relationships,’ persist long beyond the first months after a death (Hewson et al., 2023; Rees, 1971).

Neurological or psychiatric research into ADEs is even more limited (Fénelon et al., 2011; Pait et al., 2023; Reckner et al., 2020; Toh et al., 2023) . In those who have approached the topic, the studies have been often limited to single case studies, heterogenous groups with small sample sizes, and inconsistent methodologies (Kamp et al., 2020). There are, however, neurological conditions, such as PD, Alzheimer’s disease, or dementia with Lewy bodies, in which patients may experience hallucinations as part of the disease, and these hallucinations may include ADEs.

### Patients with Parkinson’s disease Having Hallucinatory Experiences

As our opening vignette suggests, PD is one clinical condition that is of interest for the study of ADEs. PD is most diagnosed as a movement disorder with accompanying motor symptoms, such as bradykinesia, rigidity, tremor, and gait disturbance (Han et al., 2018). However, nonmotor symptoms such as sleep disturbance and psychiatric conditions are becoming more clinically relevant in understanding the disease (Aarsland & Kramberger, 2015; Han et al., 2018; Thanvi et al., 2003). Approximately half of patients with PD experience hallucinations, which increase in frequency and severity as the disease progresses (Fénelon et al., 2011; Ffytche et al., 2017b). Early in the disease progression, it is more common for people with PD to experience so-called “minor” hallucinations such as PH, passage hallucinations (in which a person or animal feels to be passing by), and visual illusions (e.g., misperception of a real stimulus) (Ffytche et al., 2017a; Ravina et al., 2007). These may progress towards formed visual hallucinations, which, along with auditory hallucinations are considered “structured” hallucinations, and, recent research suggests, cognitive impairment and decline (Bernasconi et al., 2021, 2023). Hallucinations may occur repeatedly and, in some patients, occur weekly or even daily (Fénelon et al., 2000, 2011).

There are several theories as to why these experiences occur, both in the context of PD and in neurology generally. Early theories suggested that PH arise from a misattribution of our own bodily self (Brugger et al., 1996). This progressed as more neuroimaging and sensorimotor data emerged to suggest that PH experiences of neurological origin were related to altered bodily self-processes (Blanke et al., 2014). Other studies suggest that PH are hallucinations of social origin and may be related to areas in the brain associated with affective, self-related, and autobiographical memory functions (Fénelon et al., 2011; Nielsen, 2007). The social context combined with sensorimotor effects may be one way to connect PH more broadly to the phenomenon of ADEs and accounts related to attachment and continuing bonds theories.

PD most often affects elderly adults, therefore there is an age overlap in prevalence of PD and ADEs, the latter of which often occur in older adults (although younger adults have also report ADEs) ((Castelnovo et al., 2015). Individual patients with PD may experience ADEs more often and more regularly than otherwise healthy, acutely bereaved adults, and may report aspects that are less easily observed. Although hallucinations are now reported more widely among patients with PD (Bernasconi et al., 2021, 2022), there is still limited data on the prevalence and modality of such hallucinations. Some studies have examined the content or qualities of the hallucinations, such as their spatial location, affect, or if the person that is experienced in the hallucination is a known person and identified as such (Fénelon et al., 2011; Pait et al., 2023; Reckner et al., 2020; Toh et al., 2023). This latter subject of investigation relates more closely to ADEs.

### From hallucinations to ADEs

Previous studies have investigated different aspects of hallucinations in PD, especially “minor” hallucinations (Bernasconi et al., 2021; Blanke et al., 2023; Potheegadoo et al., 2021). These and other studies have found that some of these people with PD identify their hallucinations as known deceased people (Fénelon et al., 2000, 2011; Reckner et al., 2020). In the present study, the participants that report these kinds of hallucinations are drawn from a multi-center clinical study on hallucinations in PD, which included an intensive, detailed interview about the phenomenology of all patients’ experiences and hallucinations.

The aim of the case studies presented here is to provide a rich description of the phenomenology of recurrent ADEs and the way in which participants make sense of their experiences. We focused on six participants with PD who described hallucinations in which they identified a known, deceased person and maintained a fairly high level of insight. In our analysis, we asked the following main questions: what is the content of the ADE, how does the ADE evolve over time with respect to PD onset and the time from the death, and what does this identification of an ADE within a larger disease framework potentially tell us about the mechanism of the experience itself?

## Methods

To investigate the phenomenological characteristics of the ADEs in our sample of participants with PD, we examined semi-structured interview data gathered during a more wide-reaching research study on hallucinations in PD (n= 77). The study (2019-2022) included patients with PD who both did and did not report hallucinations at home. As part of a long battery of neurological tests and evaluations, patients also completed a semi-structured interview. In this paper, we will focus on the video and transcription of the clinical interview, using phenomenological and textual analysis to better understand the subjective experience of ADEs in these patients. We have employed thematic analysis to identify and interpret the patterns and themes in the qualitative data (Naeem et al., 2023).

All participants provided their written informed consent for participating in this project. The project was approved by the Geneva Cantonal Commission of Ethics in Research (CCER): Ethics protocol n° 2019-02275. Prior to the interviews, participants were given information outlining the broader study’s purpose and procedure. The interviews were both audio and video recorded and then transcribed. We obscured all identifying information, and pseudonyms are used throughout this paper.

### Research Participants

Data from six participants was included in the current sample of patients who positively identified an ADE as part of their experiences in the interview. All the participants had been diagnosed with PD prior to inclusion into the study. The mean duration of their disease (time elapsed between diagnosis and interview) was 10.7 years (range: 6-20 years; median: 8 years). The participants experienced a variety of modalities of hallucination, both minor and structured. The identity of the bereaved presence was also varied, even within participants on some occasions. Demographic, medical, and hallucinatory experience data are given in Table 1.

**Table 1.** Demographic, medical, and hallucinatory experience data for six participants.

| Name<br>(pseudo-<br>nym) | Sex | Age,<br>age<br>range | PD<br>Duration,<br>years | Identity of<br>Deceased<br>Presence | Time<br>Since<br>Death | Type of<br>hallucination | Frequency<br>of<br>Experience | Emotional<br>Experience |
| --- | --- | --- | --- | --- | --- | --- | --- | --- |
| **Colette** | F | 70-80 | 16 | Friends or family members | Unclear | Visual | 3-4x per week | Neutral |
| **Michel** | M | 50-60 | 6 | Parent | Many years, unclear | Presence | 1x per month | Wonder, incomprehension |
| **Françoise** | F | 70-80 | 7 | Husband / Parent | 20 years / unclear | Presence | 3-4x per month | Happy |
| **Alain** | M | 70-80 | 20 | Mother-in-law | ~3 years | Presence, auditory | 1x per day | Curious, comforting |
| **Monique** | F | 70-80 | 9 | Husband | 30 years | Presence, passage | 2-3x per week | Positive impact comforting |
| **Claudine** | F | 50-60 | 6 | Not specific | Unclear | Presence, auditory | 2-3x per week | Frightening |

### Data Collection and Analysis

For our analysis, we rely primarily on the transcribed interviews with participants that took place in our laboratory. The interviews lasted around one hour and included wide-ranging questions that asked about the participants’ general hallucinatory experiences. From these interviews, we narrowed down the sample to six participants who spoke about an ADE despite no specific question for this type of experience.

These data were analyzed using thematic analysis, generating specific keywords and codes from extracted quotes (Braun & Clarke, 2006; Naeem et al., 2023). We took a grounded theory approach, as the original transcripts did not ask specific questions about ADEs, so we examined the data with a careful attention to these parts of the interview in which participants voluntarily described their perceptual experience of a deceased loved one (Levitt, 2021). The keywords and codes were organized into larger themes and connected with our theoretical structure with continual reference to the source of the extracted quotations. French was the original language of the study, and we used translation software to translate the interviews into English and these generated translations were edited and refined by a research team member. These translations were reviewed, and the translations made more fluid by a second bilingual French and English researcher.

## Results

### Perceptual and Identity Aspects of ADEs

As outlined in Table 1, participants reported different types of perceptual experience and had different personal relationships to the deceased presence identified both across the sample and within individual subjects. To illustrate their diversity, we report on the type of hallucinations, the identities given to these hallucinations, and how and when they occurred. Some participants experienced ADEs of different modalities and different people. The ADEs were mostly PH. Thus, five of six participants (83%) described PH and, of these five, two (33%) also reported having ADE related to passage hallucinations (i.e., the feeling that a deceased person passes the participant rapidly on one side of the body in the immediate environment, but without seeing or hearing the deceased). Alain was one of three participants who had mixed PH and other kinds of hallucinations (i.e., auditory, passage). Alain has been diagnosed with PD for 20 years and his hallucinations occur relatively frequently. His now-deceased mother-in-law, who lived with him and his partner before her death, is described as a presence that sometimes whispers words to him. Another participant, Claudine, reported having presence ADEs sometimes combined with bodily auditory components (i.e., breathing, footsteps). Claudine hears a breath on her right shoulder and, separately, senses the presence that she describes as possibly a ghost. Colette, who is in her 70s, reported visual ADEs, and these were not accompanied by other kinds of ADEs. In these moments, Collete sees one to two people on the balcony outside of her room. She explains that “almost everyone I see has died,” and specifies that they are friends or family members. These hallucinations have persisted for 10 years, but she did not experience them before her PD diagnosis 16 years ago.

In addition to the variety of types of hallucinations that occurred between participants, the identity of the deceased presence ranged from first-order relatives like parents and spouses (57% of ADEs) to more distant relatives like mother-in-law (14%) and then to “deceased people” with unclear identities beyond being deceased (28%). Their identities were sometimes very clear and other times less distinct. Alain and Michel clearly identified their mother-in-law and parent, respectively. Françoise, who was widowed nearly 20 years ago, and Monique, also a decades-long widow, were in flux about the identity of the presence. Françoise thought it could be either her deceased husband or her deceased parent, while Monique was more confident that it was her deceased husband, but the experience was too indistinct for her to be completely sure. Both Claudine and Colette had even less specificity in their respective identifications.

Claudine’s ADEs were certainly deceased people that she knew, but she could not give them specific identities. Colette also described seeing a group of people that she recognized and who were all deceased but could not name them individually. The ambiguity in at least three of the six participants deviates with some of the existing research on ADEs in healthy participants that tend to describe the experiences as specific and evident, which we will describe further in the discussion below.

### Conditions of Occurrence

The time and context in which the participants experienced ADEs were consistent. All six participants’ ADEs occurred when they were alone and ADEs were relatively brief, four of six lasted only a few seconds. Most occurred while in their homes, except for Françoise, who reported having an ADE while walking alone in the countryside near her home. Similarly, all participants but Françoise described the ADE occurring at night or in the evening (because Françoise walks during the morning and afternoon). Of the five participants who experienced ADEs in the evening or at night, one participant described the ADE occurring in the time just before sleep while in bed. The other four participants describe themselves as standing or doing simple activities inside their home while having the experience, in full wakefulness and not related to sleep.

Michel has been diagnosed with PD for six years. In his home at night, Michel feels these shivers or chills that he attributes to his deceased parent. When they are stronger, he has an even greater sensation that it is them. He clarifies at different points that sometimes it is his parent, while other times he just thinks it is or attributes it to his parent and then other times is unsure if it’s them. Alain experiences his ADEs in his kitchen in the evening. He describes sounds of breath like a draught that he attributes to his mother-in-law and that “…where I hear it most is when I get up twice at night. I am alone in my kitchen, there isn’t anyone there, there isn’t any sound outside. And I set my table, I do a little turn to move a little bit, and it was there that I hear it.” Sometimes these breaths sound like his mother-in-law is whispering, “courage, courage.” Other times he describes these experiences more like sense of presence only.

Presence research in patients with PD is concerned with the spatiality of the experience. Our participants had varying degrees of spatial proximity to the perceived presence. Four of the six participants reported the ADE presenting one meter or less from them. Colette reported the visual ADE a few meters away on the balcony, which is a more common spatial distance for visual hallucinations. Michel experienced his presence ADE from 10 meters away, but also described it as being within the same room in his home, so this distance may be overestimated. Three participants reported the ADE as occurring behind them. Two described the ADE as next to them (one on the right, one on the left). Colette did not specify the distance or side but described the ADE as appearing on the balcony a few meters away, presumably in front or on the side of where she was lying in bed.

Critically, our participants who experienced ADEs were not acutely bereaved, meaning the people identified in their ADE had not recently died. As described in Table 1, Françoise’s husband died nearly 20 years before our conversation (her PD was diagnosed 7 years previously) and she had no ADEs around the time of his death.

Similarly, Monique’s husband died over 30 years before our conversation and more than 20 years before her PD diagnosis. She had similarly not reported having an ADE prior to her diagnosis. The other two identified ADEs—Michel’s parents and Alain’s mother-in-law—had also died several years previously. Because Colette and Claudine were not specific in their identifications for their ADEs beyond friends or family who had died, there is less specificity about their appearance relative to their death. However, neither reported a recent loss.

### Emotional characteristics

The emotional characteristics of these experiences were diverse across our sample. The general affect is positive, although participants sometimes used words or phrases that are more neutral or even negative. Overall, 67% of participants had positive feelings during and/or after the experience. These included feelings of wonder, happiness, and peace. Françoise and Monique described the ADEs as very comforting and as inspiring a sense of happiness and peace. Françoise says that she finds the ADEs so agreeable that, “in the end I like them. I don’t resent them, on the contrary, let them stay there.” Michel and Colette described having neutral responses, with Michel describing some sense of wonder at the experience but no strong emotional reaction. Alain was also neutral, although trending more positively, expressing a sense of curiosity about what was happening and that he found it comforting in some ways. As briefly mentioned above, Claudine described some fear and anxiety in perceiving these deceased people. Her experience was also the least specific, not enabling her to clearly identify any of the people whose presence she felt.

With respect to how insight relates to these affective experiences, we see a parallel in recognition of the presence and how that impacts the participants’ feelings. Perhaps because she was formerly a psychiatrist, Monique frequently assures us that she knows that her ADEs are a result of her PD. During her ADEs, she is lying in bed and feels the presence that she identifies as either a living friend or parent, or her deceased husband. Unlike some of her other hallucinatory experiences, which can feel scary or anxiety-inducing, the experiences when she identifies the presence as her deceased husband (or her living friend/mother) are positive. This might be more about known company rather than her husband specifically because, as she explains, “I feel the presence of someone reassuring, and that’s always positive.” This contrasts with the way that Claudine experiences many of her hallucinatory experiences, which are generally negative. Her ADEs are vaguer, without clear relationships, but she understands them to be deceased people that she knows. Yet, when she feels the breath behind her that signals this experience, she will sometimes try to communicate with the presence. She asks, “Who is it? What do you want?” and explains, “at first, I was afraid, but sometimes, I said to myself: But who is it? Who’s behind me? Then I say, but don’t you want to talk? Is someone there? And there’s no answer.” Then Françoise’s experience is unique in that it takes place outside. She describes walking in countryside lanes alone during which time it feels like someone is trying to overtake her on the path, but no physical person is there. At that moment she feels, “It’s peaceful. I’m peaceful. Yes…

Oh, maybe I’m saying a message to those who have left.” After she explains this moment in the interview, she reveals that she has a feeling that it the presence is her husband or parent. The identity is different at different times.

### Understanding the Experiences

A final theme that emerged was the way in which the participants made sense of or understood the ADEs. For some, the process of sense-making was cognitive, and they used language related to cognition or active thought. Others used words more related to imagination or the mind playing “tricks.” Alain was particularly methodical in his understanding of the ADE. He explained that he always looks for a logical explanation when he has the experience of sensing his mother-in-law. When asked explicitly if he thought the presence was his mother-in-law, he scoffed and said “no, she’s dead!” However, despite this assertion, he also recounts actually hearing his mother-in-law whisper “courage, courage,” but in the next reply he says it is just as likely that he invented this. Michel employs the same kind of justification or reconsideration when he experiences the feeling of his deceased mother. Monique also believed that there was always an explanation for the sensations of the ADE. These three participants used the strongest cognitive-style language when describing their ADEs. They were also the ones who specified that they held secular, agnostic, or atheist beliefs (Michel, Alain, and Monique, respectively).

Slightly distinct from this cognition or process-oriented language, the other participants used words related to imagination, subconscious, or passive thoughts. Colette described the experiences as being from her “subconscious” rather than being manifest in the world. Similarly, Françoise says that she believes that her feeling the presence of her deceased husband or father is only in her thoughts, not a reflection of reality. She agrees that she is “imagining” the deceased person to be there. Claudine alone remains completely outside cognition or thoughts-related language. She expresses real uncertainty about the experiences of ADEs, simply saying that she cannot explain why they happen, and that they are “bizarre.” Claudine was the only participant that identified as having more spiritual or religious beliefs, expressing at one point that she thought some of her hallucinations (not the ADEs) were djinns.

## Discussion

We sought to better understand how ADEs are reported in a group that has more frequent hallucinatory experiences than non-clinical populations, how these ADEs relate to onset of the death of a loved one and to PD onset, and what the ADEs from this perspective may explain about the mechanism of the experience itself. Our analysis shows that these ADEs share several important aspects with ADEs reported in acutely bereaved and/or otherwise healthy populations and show important differences. We consider the theoretical implications of ADEs in people with PD.

### Comparison with ADEs in Other Populations

There are both important similarities and differences between our group and those who have ADEs without PD. Our group of patients with both PD and ADEs and the general population of people with ADEs were close with the identified, deceased person and had, on average, a more positive experience. Like the general report of ADEs, our participants were more likely to identify the ADE as a close other (in this case, parent, in-law, or spouse) (Jahn & Spencer-Thomas, 2014; Suhail et al., 2011). However, some studies have shown that even less “close” relationships are reported during these experiences, such as grandparents or close friends (Mäkikomsi et al., 2023). Although in-laws are often not included as a “close other” for the purpose of these studies, Alain lived with his mother-in-law prior to her death and described his relationship as being close.

Participants perceived their ADEs generally as positive, particularly if we consider senses of wonder and curiosity as positive or at least not negative responses. This supports previous findings that ADEs offer comfort and are regarded as positive experiences in a Western context (Grimby, 1993; Kamp et al., 2019; Rees, 1971). The negative experiences reported by Claudine are not completely unusual, as there are some reports of negative ADEs in the literature (Kamp et al., 2019, 2020). We suggest that this experience may stem from her not fully knowing or identifying the deceased people. The negative emotions associated with this uncertainty may also suggest that the identification of an ADE as a close other is a kind of coping mechanism or social feature. The duration of the ADE was also relatively consistent as most participants reported the experience lasting between a few seconds to a maximum of five minutes. However, unlike reported ADEs in the literature in which these are often singular occurrences, the present participants had recurring experiences of the same person in a similar situation in multiple instances across time, with all six reporting these experiences as occurring with frequency over the course of the last one to three years. In this way, ADEs in PD are not singular occurrences, and may occur over several years. In addition, they may fluctuate in their identification of the deceased person, which does not often happen in ADEs in other contexts.

### Social Aspects of ADEs

Many studies have suggested that ADEs are a result or part of relationship-based interruptions, such as continuing bonds (see Hewson et al., 2023 for review) or social seeking behavior after death (White et al., 2017; White & Fessler, 2018). As expected, several social elements emerge in our observations. The participants of this study clearly identified the ADE as a close other (Jahn & Spencer-Thomas, 2014; Suhail et al., 2011). The social bond with these individuals may play a role in the way in which the manifestation of other traits, like loneliness or anxiety, shape the hallucinatory experience. Two participants spoke about how the appearance of the ADEs, and other hallucinations, were a sense of comfort and welcomed when they were feeling lonely. One participant specifically described her loneliness as part of this experience and shared her both general anxiety and specific worries about the ADEs. Although perhaps intuitive from a bereavement perspective, loneliness and social isolation have also been linked to increased frequency of hallucinations and to PD in general (Brederoo et al., 2023; Prenger et al., 2020). The present data extend these findings to ADEs in PD by highlighting the specific social phenomenology of parkinsonian ADEs. Future work should investigate how such social ‘other’-related mechanisms of ADEs might overlap with those related to ‘self’, as previous studies have linked PH in PD to sensorimotor ‘self’-related mechanisms (Blanke et al., 2014; Bernasconi et al., 2021). This may be particularly relevant given the many shared behavioral and neural mechanisms in perceiving and representing oneself and others, especially those closer and more familiar (i.e., Maister et al., 2015). Such future work should also investigate the impact of meaning-making that may account for some of the observed differences between PH versus ADE in patients with PD.

### Participants Making Sense of ADEs

As far as we can glean from their self-report, our participants are deeply reflective in thinking about the experience and rationalizing it, both when they identify the deceased presence and report on it later. For participants, ADEs are slightly different from their other PD-related hallucinations in that they require more proactive analysis than when they can easily dismiss the sensation as part of their disease or medication treatment (also found in Reckner et al., 2020). And yet, our participants maintain a great deal of cognitive insight in the sense that they can criticize and distance themselves from their perceptions. Their repeat exposure to hallucinatory experiences more broadly allows them to report a process of rationalization or reflexivity in thinking about these experiences. Furthermore, we can benefit from these experiences happening more recently compared to many studies regarding ADEs in bereaved populations, where it is more difficult to recruit and control for recent ADEs.

Previous studies have suggested that the cultural context and personal beliefs have an influence on how ADEs manifest (Exline & Wilt, 2023; Larøi et al., 2014). However, three of our participants report belief systems that are secular, atheist, or agnostic and profess that they do not believe in the appearance of spirits or ghosts. In the cultural context of Switzerland, ADEs are not necessarily precluded, but they are not part of a standard cultural milieu. None of our participants reported any changes to their beliefs or outlook because of these experiences, which has sometimes been reported, although this may be more prevalent among those who are already religious (Penberthy et al., 2023).

Michel openly reflects on how his beliefs impact the way in which he thinks about his ADEs. He initially believes that the chill he associates with his mother is part of “what I call indoctrination. From those who talked to us when we were small…that the spirits return and that are some people who protect us up there…but I think, we grew up, of course, and that we return automatically in those seconds when we have this phenomenon. We are obliged to think something, and I simply thought of my mother.” Ultimately, Michel battles with his internal understanding that religious and spiritual beliefs have been “indoctrinated” in his mind, but he also does have this sense that his mother is sometimes there with him and is worried about his situation. In a similar way, Alain describes his experiences of seeing his mother-in-law as oscillating between momentary suspension of disbelief and then rationalization. There is an initial moment of doubt or speculation, in which he considers that she might be “alerting me” and he wonders “is there something that is going to happen?” But when asked if it is real, Alain asserts, “no, she wasn’t there! She’s dead!” There is active cognitive dissonance between the initial experience and the later reconciliation, which seems to reflect an initial, impulse identification and then a secondary meaning-making moment.

### Meaning-Making in ADEs

Another key consideration in the way in which our participants made sense of ADEs are related to the time since the death of the perceived person. Interestingly, our participants tend to be many years past their acute phases of bereavement. While the identification of the close other in the ADE may indicate a continued bond, they are also actively meaning-making. We are interested in this process of sense-making for our participants because it offers a novel kind of understanding because of their repeated exposure to ADEs. In the limited number of studies about ADEs, subjects will sometimes report multiple instances of ADEs, but we have little information about the frequency within subjects in these studies (Grimby, 1993).

Our participants engage in processes of meaning-making that closely mirror those following other kinds of traumatic events. Park’s (2010) meaning-making model suggests that when a stressful event occurs (in our case, an ADE or hallucination), the person experiencing it must actively reconcile this experience with their global meaning, or it must fundamentally change. While this perspective might suggest that they could only report on the identity of the ADE if they believed it was them, we find that most of our subjects are sure that the experience is not the actual person who is deceased, in a literal sense. Instead, they have the experience without the accompanying belief that it is “real” from a spiritual or religious sense. This suggests that future studies may need to examine ADEs in the context of different mechanisms rather than those only related to belief. Although attribution is an important aspect of meaning-creation (i.e., saying “this is a person I know who died”), it is also a relatively cognitive process either quite concurrent with or just after the experience itself.

## Conclusion

Patients with PD who have recurrent hallucinatory experiences with preserved insight offer a unique opportunity for the study of ADEs. Our participants can reliably compare ADEs to other types of hallucinations and have provided key information about the nature, recurrence, and circumstances of the experience. ADE research is often limited to single instances reported months or years after the person had the experience and the lack of intra-subject reporting means it is difficult to gather data about why or when ADEs occur.

Our findings suggest that there is overlap between ADEs in patients with PD and the general report of ADEs in bereaved or otherwise healthy populations. Both groups experience these moments as short in duration, typically while alone, and as positive. The key differences, and strengths, of the experiences of patients with PD is that the ADEs are recurrent and occur outside of the acute grief period. Perhaps the most telling, however, is the momentary lapse in meaning reconciliation, where our participants have a perception that then results in identification or not. Unlike some kinds of hallucinatory experiences that are more vague or unclear, there seems to be active language of thinking about, wishing, and otherwise constructing beliefs around the identity of the ADE. These moments of meaning-making may involve a social component to the hallucinatory experience that has not been deeply explored previously. The alteration in social perception found in patients with PD and hallucinations (i.e., Albert et al., 2024) may thus give rise to hallucinations with social content (e.g., identified, deceased loved ones versus unknown people). Additional work is also needed to investigate self-related sensorimotor mechanisms in ADEs in PD and beyond. Future research may aim to test bereaved individuals using behavioral and neurocognitive measures alongside phenomenological methods described here to get a multi-faceted view of ADEs as a complex phenomenon.

## Data Availability Statement

The data that support the findings of this study are available from the corresponding author, C.A., upon reasonable request.

